# Brain activity is contingent on neuropsychological function in an fMRI study of Verbal Working Memory in Amyotrophic Lateral Sclerosis

**DOI:** 10.1101/2021.01.04.21249202

**Authors:** Xenia Kobeleva, Judith Machts, Maria Veit, Stefan Vielhaber, Susanne Petri, Mircea Ariel Schoenfeld

## Abstract

Amyotrophic lateral sclerosis (ALS) is a devastating neurodegenerative disease that causes progressive degeneration of neurons in motor and non-motor regions, affecting multiple cognitive domains. To contribute to the growing research field that employs structural and functional neuroimaging to investigate the effect of ALS on different working memory components, we conducted a functional magnetic resonance imaging (fMRI) study exploring the localization and intensity of alterations in neural activity. Being the first study to specifically address verbal working memory via fMRI in the context of ALS, we employed the verbal n-back task with 0-back and 2-back conditions. Despite ALS patients showing unimpaired accuracies (p = 0.724) and reaction times (p = 0.0785), there was significantly increased brain activity of frontotemporal and parietal regions in the 2-back minus 0-back contrast in patients compared to controls using nonparametric statistics with 5000 permutations and a T-threshold of 2.5. This increased brain activity during working memory performance was largely associated with better neuropsychological function within the ALS group, suggesting a compensatory effect. This study therefore adds to the current knowledge on neural correlates of working memory in ALS and contributes to a more nuanced understanding of hyperactivity during cognitive processes in fMRI studies of ALS.

## Introduction

Amyotrophic lateral sclerosis (ALS) is characterized by progressive impairment of motor and non-motor brain regions. Reflecting the increasing scientific attention on non-motor effects of ALS ^1–7^, current epidemiological findings indicate that up to 55 % of ALS patients exhibit cognitive deficits, including up to 20% of patients with characteristics of frontotemporal dementia ^8–12^. These deficits mostly manifest in impaired verbal fluency and executive dysfunction ^13^, but also in other cognitive domains, such as language, social cognition, and different components of memory ^14,15^. Given that cognitive and behavioral deficits are associated with poor survival in ALS ^16,17^, better understanding of these deficits is necessary for development of therapeutic strategies.

One important component of memory, that has been considered in ALS research, is working memory. It is the ability to temporarily store information during cognitive processing ^18^. If impaired, it causes general higher-order cognition to deteriorate. Based on Baddeley & Hitch’s seminal paper from 1974 ^19–21^, the current dominant model of working memory comprises four principal components: Two unimodal storage systems (i.e., the phonological loop for verbal and numerical information and the visuospatial sketchpad for visual information), the episodic buffer (storing multimodal information in an episodic representation), and the central executive (which controls these storage systems and manipulates information) ^22^.

Prior research has assessed the impact of ALS on working memory using behavioral and functional imaging measures during the execution of working memory tasks. Most research focused on the patients’ working memory capacity. Such studies utilized tasks that either address the phonological loop or the visuospatial sketchpad, with the majority of studies showing interest in the former. Specifically, studies addressing the phonological loop mostly employed variations of the digit span task as measures of working memory capacity, which was mostly found to be unimpaired in ALS ^13,14,23–25^. Research on the visuospatial sketchpad employed visuospatial sequence tasks and more complex tasks (such as the visual 2-back task) and yielded mostly comparable or marginally weaker behavioral performance results for ALS patients relative to controls ^25–27^.

In contrast to comparisons of neuropsychological performance, studies have placed much less attention on brain activity alterations in ALS patients. In fact, employing event-related potentials or functional magnetic resonance imaging (fMRI) during working memory task execution have revealed localized abnormal frontoparietal brain activity patterns in ALS patients. This suggests that the behavioral parity to control groups in ALS patients is accompanied by compensatory or potentially disease-related brain network changes.

For example, Hammer et al. ^25^ applied event-related potentials and found parietal differences between the storage of figural and spatial information during a visual 2-back task in ALS patients, which were not present in healthy controls. Relatedly, Vellage et al. ^27^ used a visuospatial working memory task with different levels of memory load and distractors, localizing neural activity related to working memory capacity via fMRI. They found significantly different levels of frontal brain activity in ALS patients compared to controls. Specifically, they reported lower frontal brain activity during storage of working memory content (arguing that this can be attributed to a breakdown of storage capacity), while higher frontal brain activity was observed during filtering of relevant stimuli (arguing that this can be attributed to compensatory hyperactivity).

In prior literature working memory in ALS, significantly more research has addressed the phonological loop than the visuospatial sketchpad ^13,14,23–25^. Given this proportion, it is remarkable that none of the studies addressing the phonological loop have employed neuroimaging methods or complex tasks with active manipulation of working memory content. Such complex tasks could feature a higher sensitivity to working memory dysfunction in ALS. Furthermore, using fMRI could substantiate possible working memory dysfunction and compensatory processes in ALS by describing ALS-associated patterns of neural activity involved in verbal working memory function.

In this study, we therefore sought to extend evidence on ALS-induced verbal working memory dysfunction. Specifically, by using fMRI, we investigated the underlying brain activity in ALS patients (and controls) during execution of the well-established and robust verbal n-back task ^28^, which addresses both the central executive as well as the phonological loop. By doing so, we acknowledge that such a complex task requires more active manipulation of working memory content and could be more sensitive in detecting working memory dysfunction in ALS ^25^. Building on prior research, we hypothesized comparable or subtly diminished behavioral accuracy associated with ALS. We complement previous studies that focused on visuospatial working memory; in consequence, we aim at supplying information on whether potentially impaired working memory performance and accompanying brain hyperactivity levels occur in both the phonological loop and the visuospatial sketchpad in patients with ALS. We expected ALS patients to show an increased activity of frontoparietal brain areas associated with reduced executive function. We especially aimed to address the potential interaction of neuropsychological function that is known to be impaired in ALS patients (executive function, verbal fluency and language function) with working memory performance and observed brain activity.

## Materials and Methods

### Subjects

28 ALS patients were recruited from the ALS clinics of Hannover Medical School and University of Magdeburg. They had been diagnosed by a board-certified neurologist according to the revised El Escorial criteria (i.e., diagnosis of possible, probable, laboratory supported-probable or definite ALS ^29^) and did not show clinical signs of other current neuropsychiatric disorders — especially no dementia according to the ALS-FTD criteria ^30^. They were all able to press a button with their right hand and lie horizontally in the MRI scanner. 21 controls matched for age, gender, and education and without a history of neuropsychiatric disorder participated in the study. Subjects were excluded if their task performance during a practice run was insufficient (< 2/3 of correct responses) or if they exhibited intolerable head movement (see description of preprocessing below). This led to the exclusion of four patients due to excessive motion and two patients due to insufficient task performance, resulting in 22 ALS patients included in the analysis.

Functional impairment was classified by the revised version of the ALS functional rating scale (ALSFRS-R ^31^), and the severity of motor symptoms was assessed by a standard neurological examination. The neuropsychological evaluation consisted of the cognitive part of the Edinburgh Cognitive and Behavioural ALS Screen (ECAS, German version) ^32^. In addition to analyzing the group means, the ECAS was used to categorize ALS patients into subgroups according to the revised Strong criteria ^33^: ALS-ci (cognitively impaired in at least one ALS-specific cognitive domain) and ALS-ni (no cognitive impairment) according to the ECAS cut-off values ^34^.

The study was approved by the ethics’ committee of the medical faculty of the University of Magdeburg and all subjects provided informed and written consent.

### Experimental Design

Working memory function was assessed using a letter version of the 2-back task ^35^. This task was part of a bigger (yet unpublished) fMRI study. The task was presented with the NBS Presentation software. Before entering the MRI scanner, all participants performed a short practice trial.

During the experiment, each condition started with a 5-second “instruction” condition. In the 0-back condition “A”, 15 capital letters were presented in a random order. The participants were asked to press a button with their right index finger whenever the letter “N” appeared on the screen (in four out of the 15 trials). During the 2-back condition “B”, participants were required to press the button if the presented letter was the same as the one that appeared two items before (in four out of 15 trials in a pseudorandom order to avoid successive occurrence of target trials). The 0-and 2-back task conditions were presented for 400ms, followed by an inter-stimulus-interval of 1.6 seconds, resulting in four blocks of 30seconds, divided by 15 seconds of fixation and five seconds of instruction. The order of the two conditions was ABBABAAB for all subjects.

### Analysis of Neuropsychological and Behavioral Data

Reaction times and error rates recorded for each participant. Behavioral and neuropsychological data were compared between groups using (M)ANCOVAs via SPSS Statistics and JAMOVI ^36^. We calculated the association between accuracy of the 2-condition of the n-back task and disease duration and ALSFRS-R in ALS using Spearman correlations. Furthermore, we evaluated the effect of group membership (patients vs controls) on accuracy and reaction times using a repeated-measures ANCOVA with covariates ECAS sub-scores for executive function, verbal fluency, and language function, and included age and gender as covariates of no interest.

### MRI Data Acquisition

Subjects were scanned in a 3T whole body MR scanner (Siemens Magnetom Verio syngo MR B19, Erlangen, Germany) using a 32-channel head coil receiver. A gradient echo single shot EPI sequence (with 42 slices, TE= 30ms, TR= 2500ms, flip-angle 80°, in-plane resolution 64×64mm, no gap, FoV 224×224mm², voxel size 2×2×3mm³) was used to acquire the functional images in an odd-even interleaved sequence (166 volumes). A T1-weighted image was acquired as well (MP-RAGE, 96 sagittal slices, thickness= 2mm, FoV 256 × 256mm, no gap, voxel size 1×1×2mm³, TR= 1660ms, TE= 5.05ms, TI= 1100ms).

### MRI Data Preprocessing

The fMRI data was preprocessed using FEAT Version 6.00, part of FSL (FMRIB’s Software Library) with a standard pipeline with motion correction ^37^; non-brain removal ^38^; spatial smoothing (8mm FWHM); grand-mean intensity normalization; high-pass temporal filtering (sigma= 30.0s). ICA-based exploratory data analysis was carried out with MELODIC ^39^ to investigate the possible presence of unexpected artefacts or activity. Outliers were detected by observing the motion parameters from the realignment step (threshold of 0.5 mm/TR or 1.9 standard deviations away from mean). If more than 1/3 of a run’s data points were outliers, the subject was excluded from the analysis.

### First-Level Analysis

All three conditions (i.e., the instruction, the 0-back and 2-back condition) were entered into a general linear model using a boxcar function convolved with a standard hemodynamic response function. For the first-level analysis, the contrast for working memory function was calculated for the 2-back minus 0-back condition.

### Second-Level Analysis

The resulting 2-back minus 0-back contrast was submitted to group statistics and regression analyses with covariates of interest using permutation-based nonparametric statistics (FSL Randomise; http://fsl.fmrib.ox.ac.uk/fsl/fslwiki/Randomise). The analyses featured 5,000 permutations; values above a voxel-wise T-value of 2.5 (uncorrected) were considered significant. Age and gender were demeaned and included as covariates of no interest. Given the comparable means and low variance of years of education in both groups, education was not added as an additional covariate. Group means and group contrasts were calculated to compare the ALS group with controls. The association of the covariates with brain activity was examined using a regression analysis with 5,000 permutations in FSL Randomise using a voxel-wise threshold of a T-value > 2.5 (uncorrected). In the ALS group, the association of ALSFRS-R and disease duration with brain activity was examined. The association of the covariates ECAS executive, ECAS language and ECAS verbal fluency was compared between ALS patients and controls. The FSL Randomise output shows the regions that contain significant group differences in slopes between the dependent and independent variables, but it does not display the direction of the slopes. To disentangle the direction of the group differences (i.e., whether the differences were caused by a positive relation of higher neuropsychological performance on brain activity in ALS patients or by a negative relation in controls), the regression analysis’ group means were calculated as well, and were subsequently overlaid with the group contrasts. Given the small sample size (due to the rarity of the disease and challenges of ALS patients to participate in fMRI studies), we were not able to apply more conservative statistical thresholds such as FDR or FWE corrections. The resulting statistical maps were visualized with Nilearn ^40^.

## Results

### Subject Characteristics

Both ALS patients and controls were comparable in age, gender, and years of education (all p > 0.05, see Table 1). 10 patients were diagnosed with possible ALS, five with probable ALS, and seven with definitive ALS ^29^. Furthermore, 10 ALS patients fulfilled the criteria for ALS-ci, according to the modified Strong criteria ^33^. Patients’ characteristics can be found in Table 1.

**Table 1.** Means and standard deviations (S.D.) of demographical data of ALS patients and controls and clinical characteristics of the patients. There were no significant differences in demographical data (age, gender, education) between groups.

|  | Controls |  | ALS patients |  | p-value |
| --- | --- | --- | --- | --- | --- |
|  | N = 21 |  | N = 22 |  |  |
|  | Mean | S.D. | Mean | S.D. |  |
| Age (years) | 63.45 | 8.15 | 62.64 | 9.06 | 0.759 |
| Gender (W:M) | 10:11 |  | 10:12 |  | 0.887 |
| Education (years) | 14.79 | 2.07 | 15.03 | 3.14 | 0.774 |
| Site of onset (limb:bulbar) |  |  | 14:8 |  |  |
| ALS phenotype (classical: flail limb) |  |  | 21:1 |  |  |
| Disease duration (months) |  |  | 22.60 | 21.56 |  |
| ALSFRS-R |  |  | 37.60 | 4.97 |  |
| genetic/sporadic ALS |  |  | 1x c9orf72 |  |  |
| ALS-ni/ALS-ci |  |  | 12:10 |  |  |

### Neuropsychological Testing

Means and standard deviations of the neuropsychological assessments are reported in Table 2. Controls had normal ECAS subscores. The MANCOVA showed a significant effect of group (F = 2.625, p = 0.041) and age (F = 3.311, p = 0.015) but not of gender (F = 1.480, p = 0.222). Post-hoc ANOVAS showed that the two groups were not different — neither in the overall ECAS score, nor in the subdomains memory function, visuospatial functions, verbal fluency, or executive function (each p > 0.05). However, at first glance patients performed significantly worse than controls in the language domain (F = 5.938, p = 0.019). After a Bonferroni-correction for multiple comparisons, neither of these values remained significant. Despite comparable mean group values in ALS and controls, there were 10 ALS-ci and 12 ALS-ni patients within the ALS group, according to the modified Strong criteria ^33^. Due to small group sizes of the ALS-ci and ALS-ni group, we did not use these subgroups for further analyses, but instead resorted to regression analyses of the ECAS subdomains (executive function, verbal fluency, and language function) on behavioral performance and brain activity.

**Table 2.** Means and standard deviations (S.D.) of the neuropsychological screening using ECAS (Edinburgh Cognitive and Behavioural ALS Screen) in patients and controls. ANOVAS were used for comparison (here, uncorrected values are presented).

|  | Controls |  | ALS patients |  | p-value |
| --- | --- | --- | --- | --- | --- |
|  | N = 21 |  | N = 22 |  |  |
|  | Mean | S.D. | Mean | S.D. |  |
| Memory (max 24) | 17.10 | 2.86 | 15.62 | 3.98 | 0.155 |
| Visuospatial (max 12) | 11.71 | 0.56 | 11.86 | 0.35 | 0.277 |
| ALS-Non-specific functions (max 36) | 28.76 | 2.98 | 27.18 | 4.00 | 0.140 |
| Language (max 28) | 27.76 | 0.63 | 26.59 | 1.99 | **0.019** |
| Verbal fluency (max 24) | 19.52 | 4.05 | 17.64 | 5.65 | 0.249 |
| Executive function (max 48) | 38.76 | 2.36 | 38.77 | 6.23 | 0.944 |
| ALS-Specific functions (max 100) | 86.05 | 5.76 | 83.00 | 11.78 | 0.272 |
| ECAS total score (max 136) | 110.90 | 13.56 | 114.00 | 7.28 | 0.155 |

### Behavioral Performance

Accuracies and reaction times of the n-back task are reported in Supplementary Table S1. The repeated-measures ANCOVA of accuracies revealed significant effects of age (F = 8.013, p = 0.007) and condition (F = 4.701, p = 0.036), but no significant effects of gender (F = 1.162, p = 0.288) or group (F = 0.127, p = 0.724). With respect to reaction times, there was no significant effect of condition (F = 0.005, p = 0.942), age (F = 0.667, p = 0.419) or group (F = 3.264, p = 0.0785) but a significant effect of gender (F = 6.554, p = 0.014).

Neuropsychological function (i.e., executive function, verbal fluency, and language function) was found to have no significant effect on the accuracy (all F-values below 0.548 and p-values above 0.464). Regarding reaction times, there was a trend towards a positive relation between executive function and the reaction times (F = 3.203, p = 0.083, other F-values below 0.013 and p-values above 0.941). In the ALS group, there was no significant correlation between ALSFRS-R scores or disease duration, accuracy rates or reaction times (p-values above 0.198).

### fMRI Results

To assess the neural correlates of working memory execution in ALS and its alterations in comparisons to controls, we calculated group means and group differences of the 2-back minus 0-back contrast (see Figure 1). Group means of ALS patients and controls exhibited comparable frontoparietal and temporal activity for the 2-back minus 0-back contrast (peak coordinates of the activation clusters are detailed in Table S2). Between-group activity differences were observed for the 2-back minus 0-back contrast in frontotemporal and parietal regions with ALS patients exhibiting higher activity than controls. Besides increased activity, regions with decreased activity were found in the ALS group within the inferior frontal cortex, frontal pole, cerebellum and caudate nucleus relative to controls for the 2-back minus 0-back contrast.

**Figure 1.**
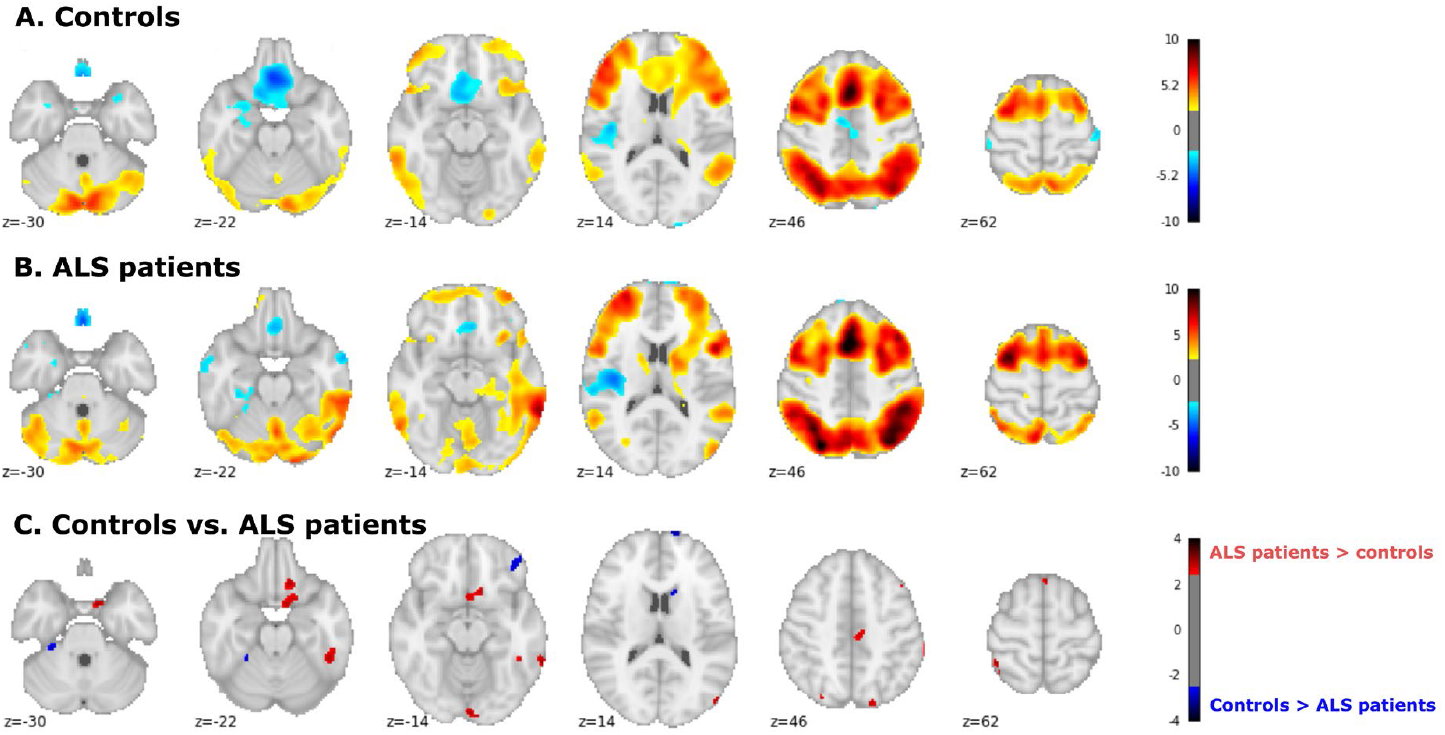
Brain activity during 2-back > 0-back contrast. Brain activity as computed as a group means of controls (A) and ALS patients (B) and group contrasts of controls vs. ALS patients (C, with brain activity in ALS patients > controls in red and brain activity in controls > ALS patients in blue). Computed by FSL Randomise, 5000 permutations, thresholded at T > 2.5.

We were then interested in associating separate contribution of the components of the working memory (phonological loop, central executive) to the observed differences in fMRI activity. To do so, we performed a regression analysis with the brain activity as a dependent variable and ECAS subdomains (i.e., executive function, verbal fluency, and language function) as independent variables. Figure 2 visualizes the group differences as they were identified by the regression analyses of the ECAS subdomains with brain activity in the 2-back minus 0-back contrast. As the group differences do not give any information on whether the differences were caused by a positive influence of higher ECAS scores on brain activity in ALS patients or by a negative influence of higher ECAS score on brain activity in controls, we performed the regression analyses for each group and neuropsychological subdomain separately and overlaid the activity maps onto the group contrasts (see supplementary Figure S1).

**Figure 2.**
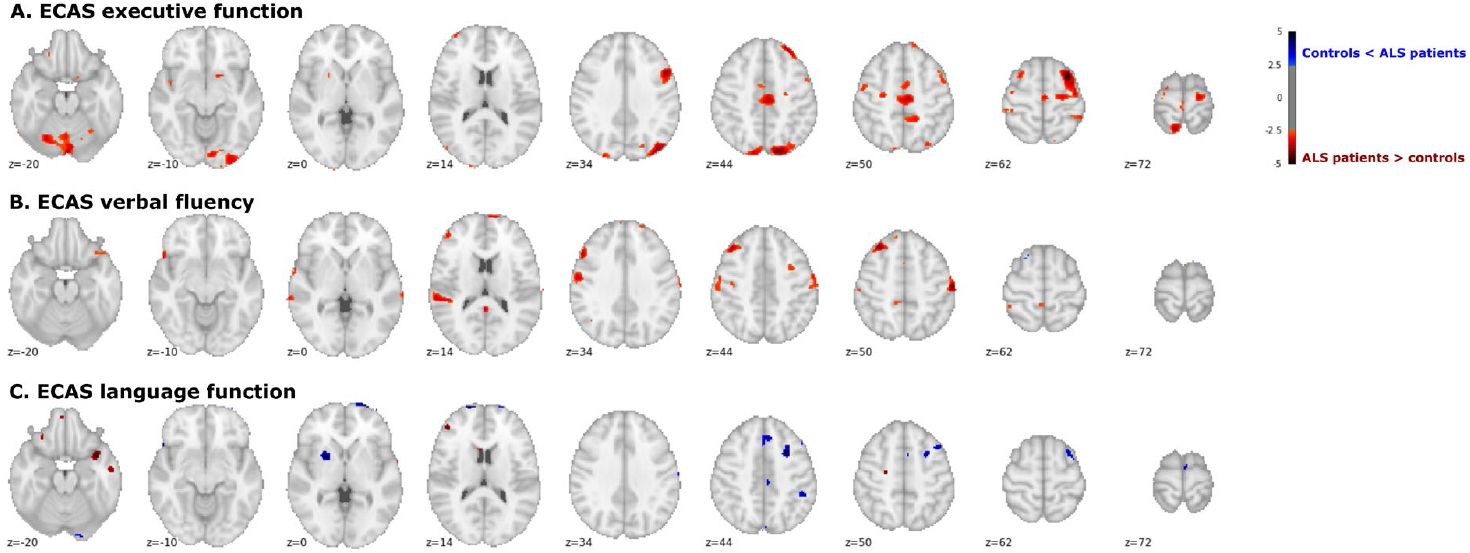
Group comparisons of the association of brain activity during 2-back > 0-back contrast with neuropsychological function. Group comparisons of regression analyses of the ECAS subdomains executive function (A), verbal fluency (B) and language function (C) on brain activity in the 2-back minus 0-back contrast. Computed by FSL Randomise, 5000 permutations, thresholded at T > 2.5).

As can be seen in Figure S1, group differences were mostly caused by negative associations of higher neuropsychological function and brain activity in controls. In ALS patients, this association was either absent or even reversed (i.e., a positive association of higher neuropsychological function and brain activity). Regarding executive function, in controls there was a negative association between higher executive function and lower brain activity of frontoparietal and cerebellar regions. In ALS patients, we found a positive association with higher executive function and cerebellar activity (which only partly overlapped with the areas where the group differences were found).

Regarding verbal fluency, we found a negative association between verbal fluency performance and brain activity in frontotemporal regions in controls, while there was a positive association between verbal fluency performance and brain activity in frontotemporoparietal areas in ALS patients, with both effects contributing to the group differences.

Regarding language function, there was a negative association between language function and lower brain activity in temporal areas in controls and a positive association between language function and brain activity in the superior frontal cortex. In ALS patients, there was a negative association between language function and brain activity in superior frontal areas and a positive association between language function and brain activity in temporal areas.

With respect to an association of clinical variables with brain activity in ALS, regression analyses revealed increased frontal and cerebellar activity as a function of higher ALSFRS-R values (Figure 3) and increased activity within the left temporal cortex as a function of longer disease duration.

**Figure 3.**
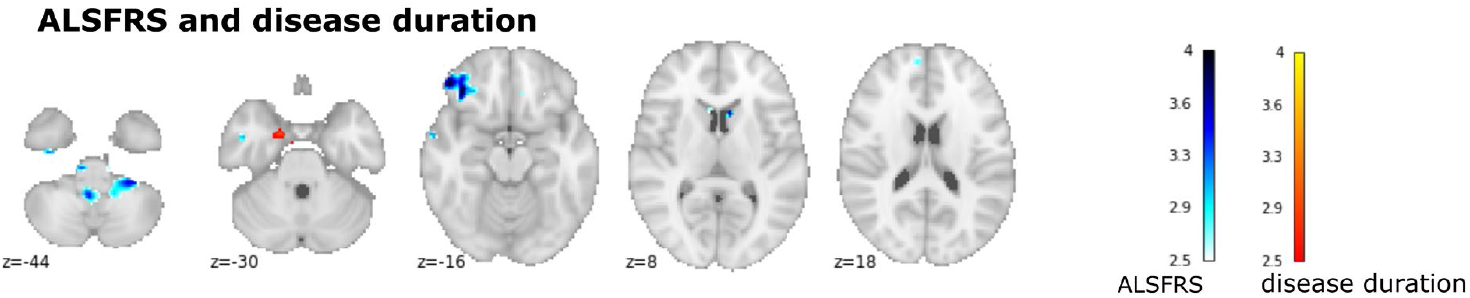
Interaction of brain activity during 2-back > 0-back contrast with clinical variables. Regression of ALSFRS-R (in red-yellow) and disease duration (in blue) on brain activity in ALS patients (computed by FSL Randomise, 5000 permutations, thresholded at T > 2.5). The ALSFRS was positive associated with brain activity of frontocerebellar regions and disease duration was positively associated with brain activity of the left temporal cortex.

## Discussion

In this study, we investigated the neural correlates of verbal working memory deficits in ALS patients. Controls and patients performed a 2-back task, and fMRI activity was associated with neuropsychological function. Patients and controls achieved overall similar behavioral measures such as accuracies and reaction times in the task. Functional imaging revealed, besides a widely comparable frontoparietal brain activity in both groups, a significantly increased brain activity of several frontotemporal and parietal regions in the 2-back minus 0-back contrast in ALS patients. Moreover, frontoparietal brain activity was influenced by neuropsychological performance in a divergent manner in controls and patients. In general, there was a negative association of superior neuropsychological function and brain activity in controls, but a positive association of higher neuropsychological function with brain activity in ALS patients. This indicates a compensatory effect in ALS: While higher executive function was largely associated with increased brain activity of frontoparietal and cerebellar regions, better verbal fluency was associated with increased activity of frontotemporal areas, and higher language function was associated with increased activity of temporal areas (exclusively in ALS patients). In addition, we found that disease severity was positively related to decreased activity within the cerebellum and left inferior frontal cortex in ALS patients, possibly indicating a breakdown of these brain resources in more advanced stages of ALS.

In accordance with previous literature ^11,13,25^, about 45% of ALS patients participating in this study were classified as ALS-ci according to the modified Strong criteria ^33^, despite similar neuropsychological test scores at the group level (except for diminished language function in the ALS group). Regarding behavioral performance, ALS patients achieved similar task accuracies and reaction times as controls. When investigating the influence of neuropsychological performance (such as executive function, verbal fluency, or language function) on behavioral performance, there was no association between neuropsychological function and task accuracy or reaction times. This argues in favor of an overall good behavioral performance of ALS patients in working memory tasks, even for verbal stimuli.

In contrast to the overall intact behavioral performance of the ALS patients, we observed some brain regions with enhanced brain activity in ALS patients, compared to controls. Both groups engaged mostly frontoparietal regions, which is in line with previous fMRI studies of working memory ^41–43^. Yet, ALS patients exhibited higher activity of frontoparietal and temporal regions in the 2-back condition. To better explain these findings, we assessed interactions between group and neuropsychological function as well as effects of disease severity and disease duration on brain activity. This approach identified several interesting insights: Overall, controls showed negative associations between higher executive function and frontoparietal activity, between superior verbal fluency performance and frontotemporal brain activity, and between language function and temporal brain activity. There was only one positive association between higher language function and brain activity within superior frontal cortex in controls. In ALS patients, these modulations of frontoparietal and temporal activity by neuropsychological function were reversed, either featuring a positive association with better neuropsychological function with brain activity, or absent association of neuropsychological function with brain activity. While in healthy subjects higher cognitive performance is usually related to less brain activity (as a sign of decreased effort ^44,45^), the relationship between cognitive performance and brain activity in ALS was found to be the opposite. This may suggest that given sufficient cognitive resources, enhanced brain activity, might compensate for disease-related effects in those ALS patients who are able to maintain a sufficient cognitive performance. We observed less brain activity in ALS patients with lower cognitive function, possibly indicating a breakdown of resources, which might probably cause further behavioral deficits in even more demanding tasks. Similar to what has been observed in patients with Mild Cognitive Impairment ^46^, we believe that changes of neural activity during cognitive processes should be regarded as dysfunctional even in absence of a behavioral deficit. This is because increased neural activity leads to enhanced energy expenditure ^47^. In our view, increased neural activity during cognitive processes is especially relevant for patients with ALS, as neurons of ALS patients might be especially vulnerable to energetic stress ^48^.

Our findings therefore suggest that whether an ALS patient exhibits hyper- or hypoactivity during cognitive processes is contingent on the cognitive resources available to that individual. Successful compensation in ALS patients might be associated with hyperactivity, and a breakdown of cognitive resources might be mirrored by hypoactivity. This finding extends current knowledge on neural correlates of verbal working memory in ALS. It also contributes to a more nuanced understanding to the common finding of hyperactivity during cognitive processes in fMRI studies of ALS ^6,49–51^. Several studies of patients with neurodegenerative diseases showed an inverted U-shaped relationship of hyperactivity and neural decline during cognitive tasks ^52,53^. Our observation of fMRI hyperactivity in patients with good neuropsychological performance and decreased activity in patients with decreased neuropsychological performance supports this body of literature on compensation.

As these associations between neuropsychological performance and brain activity were found for the 2-back minus 0-back contrast, one can suppose that these alterations are more than baseline disease-accompanying processes and contain information about functional changes of cognitive processes in ALS patients. As most differences were associated executive function and verbal fluency, it can be assumed that observed patterns of brain activity during working memory execution are mostly related to early impairments in executive function and probably to a lesser extent to language function. In consequence we suggest that the central executive is primarily affected with a relative preserving of the phonological loop function, which is well in line with previous studies ^23,54^. The characteristics of the impairment of the central executive are in accordance with several reports of reduced executive function in ALS patients ^54–56^. These results are consistent with a previous study on visual working memory describing increased frontal activity during filtering of relevant information ^27^. Besides replicating findings from this specific study, the current results contribute to the research of cognitive dysfunction in ALS in general by providing a better understanding of executive dysfunction in the context of working memory in this disease.

Besides associations of neuropsychological performance and brain activity, we found a negative association of disease severity, i.e., a lower ALSFRS-R, and brain activity within inferior frontal and cerebellar regions. This supports the notion that ALS patients, who are more severely affected by the disease, have less brain resources available for cognitive tasks. Longer disease duration was associated with increased brain activity only in a small temporal area, possibly due to a selection bias of ALS patients with a more benign disease course, who were able to participate in our fMRI study.

Interestingly, those brain areas affected and either not showing a decrease in association with higher cognitive function at all (as in controls) or even showing an increase of activity, are consistent with areas that were described in a recent postmortem neuropathological study. This study associated TDP-43 accumulation with neuropsychological function and showed associations of executive function with frontal accumulations and verbal fluency with frontotemporal accumulations ^57^. Therefore, functional neuroimaging of cognitive function in ALS might show areas of neural dysfunction reproducing known patterns of TDP-43 accumulations. In particular, our findings and their association with disease severity are in line with several previous neuroimaging studies that demonstrated a progressive disease spreading from motor to non-motor areas ^58,59^. Our study adds in-vivo neuroimaging evidence to the proposed staging scheme of ALS-associated neuropathology. Future studies dealing with task-related and resting-state fMRI should not only relate the observed neural changes to neuropsychological scores, but also specifically evaluate the observed patterns as putative biomarkers of disease progression.

Like most studies on rare diseases such as ALS, sample characteristics (i.e., small sample size and clinical phenotype heterogeneity) were determined by restricted patient availability; as such, the reported findings above should be interpreted with typical caution. Moreover, impaired respiratory and motor function have limited the number of patients who were able to participate in the fMRI task, especially in more advanced stages of the disease. To increase statistical power of fMRI studies in ALS, results of our and other research groups’ studies could be used for neuroimaging meta-analyses ^60^. To test performance in more severely impaired patients with ALS, future studies could employ longitudinal measures of cognitive function or evaluate options, e.g. by assessing of cognition using eye-tracking devices and applying simultaneous EEG to measure neural correlates ^6,61–63^. Regarding the contribution of cognitive impairment on working memory, larger multi-site studies could enable the analysis of subgroups (i.e., ALS-ci versus ALS-ni) to understand interindividual differences in cognition and behavior ^64^. Furthermore, comprehensive neuropsychological batteries for assessment of cognition might be more sensitive to detect subtle behavioral and neuropsychological impairment in ALS patients ^33,65^.

In summary, our study found comparable behavioral performance but increased frontoparietal and temporal activity during a verbal working memory task in patients with ALS. Furthermore, increased activity of working memory-related areas in association with higher neuropsychological performance in ALS patients most likely reflect compensatory effects.

## Data Availability

The participants have not consented to data sharing, therefore the individual data is not available.

## Acknowledgements

JM is funded by the federal state of Saxony-Anhalt and the European Regional Development Fund (ERDF) in the Center for Behavioral Brain Sciences (CBBS, ZS/2016/04/78113). SP is funded by the German Neuromuscular Society and Federal Ministry of Education and Research (BMBF). We thank Christian Stoppel and Christian Merkel for technical assistance with the experiment and analysis pipeline.

## Data availability

The data that support the findings of this study are available on request from the corresponding author. The data are not publicly available due to privacy or ethical restrictions. The data in this paper contain identifying patient and healthy volunteer information, which is unsuitable for public deposition. The participants of this study also did not consent to the sharing of their data.

## Supplementary material

**Figure S1.**
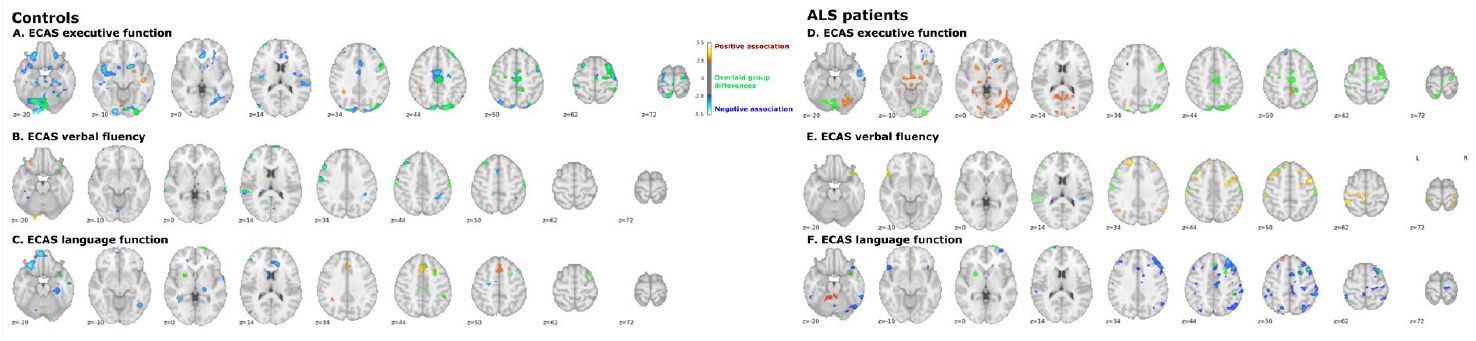
Group means of the interaction of brain activity and neuropsychological function during the 2-back minus 0-back contrast. Group means of the regression analyses of the ECAS subdomains in controls (A-C) and ALS patients (D-F) on brain activity in the 2-back minus 0-back contrast with positive (in red) and negative associations (in blue). Additional display of the group contrasts ALS patients vs. controls (as seen in Figure 2).

**Table S1.** **Means and standard deviations (S.D.) of reaction times and accuracies of the 2-of patients and controls in the 0-back and 2-back condition of the n-back-task**.

|  | Healthy controls |  | ALS patients |  |
| --- | --- | --- | --- | --- |
|  | N = 21 |  | N = 22 |  |
|  | Mean | S.D. | Mean | S.D. |
| 0-back condition |  |  |  |  |
| Accuracy (in %) | 99.20 | 2.18 | 98.86 | 2.47 |
| Reaction times (in ms) | 494.71 | 37.10 | 547.68 | 73.03 |
| 2-back condition |  |  |  |  |
| Accuracy (in %) | 89.20 | 4.036 | 90.30 | 6.31 |
| Reaction times (in ms) | 656.07 | 84.42 | 677.58 | 128.70 |

**Table S2.**
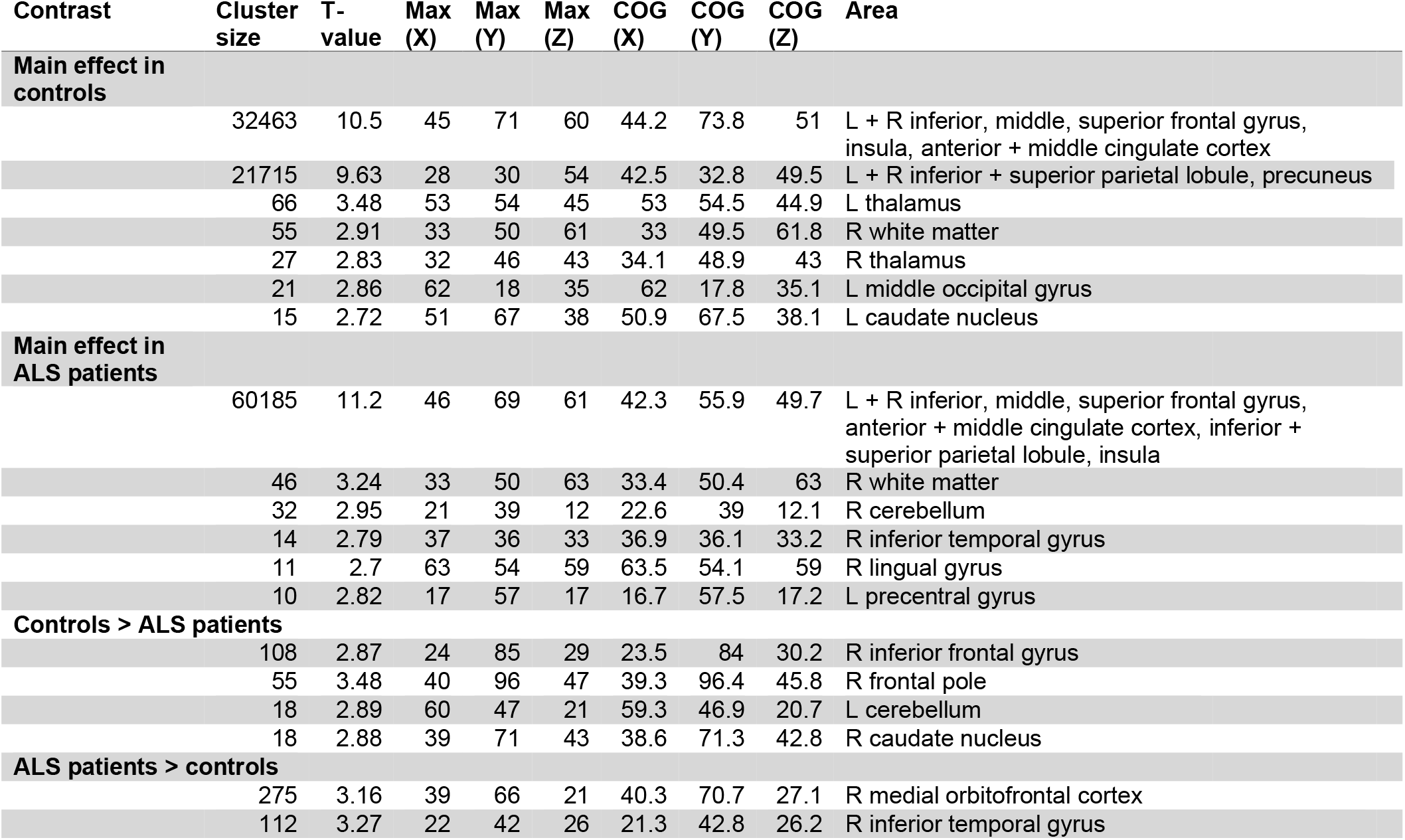
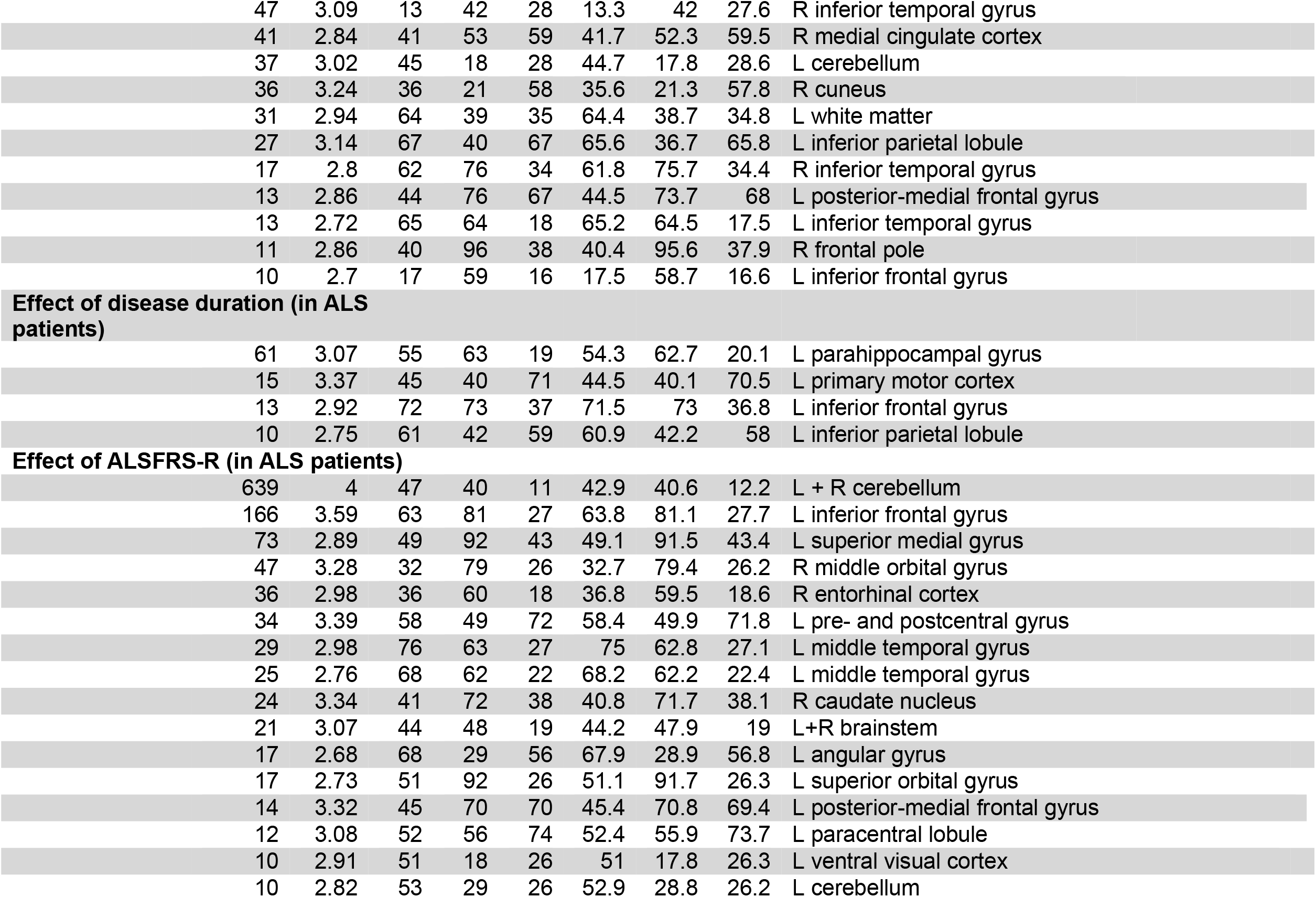
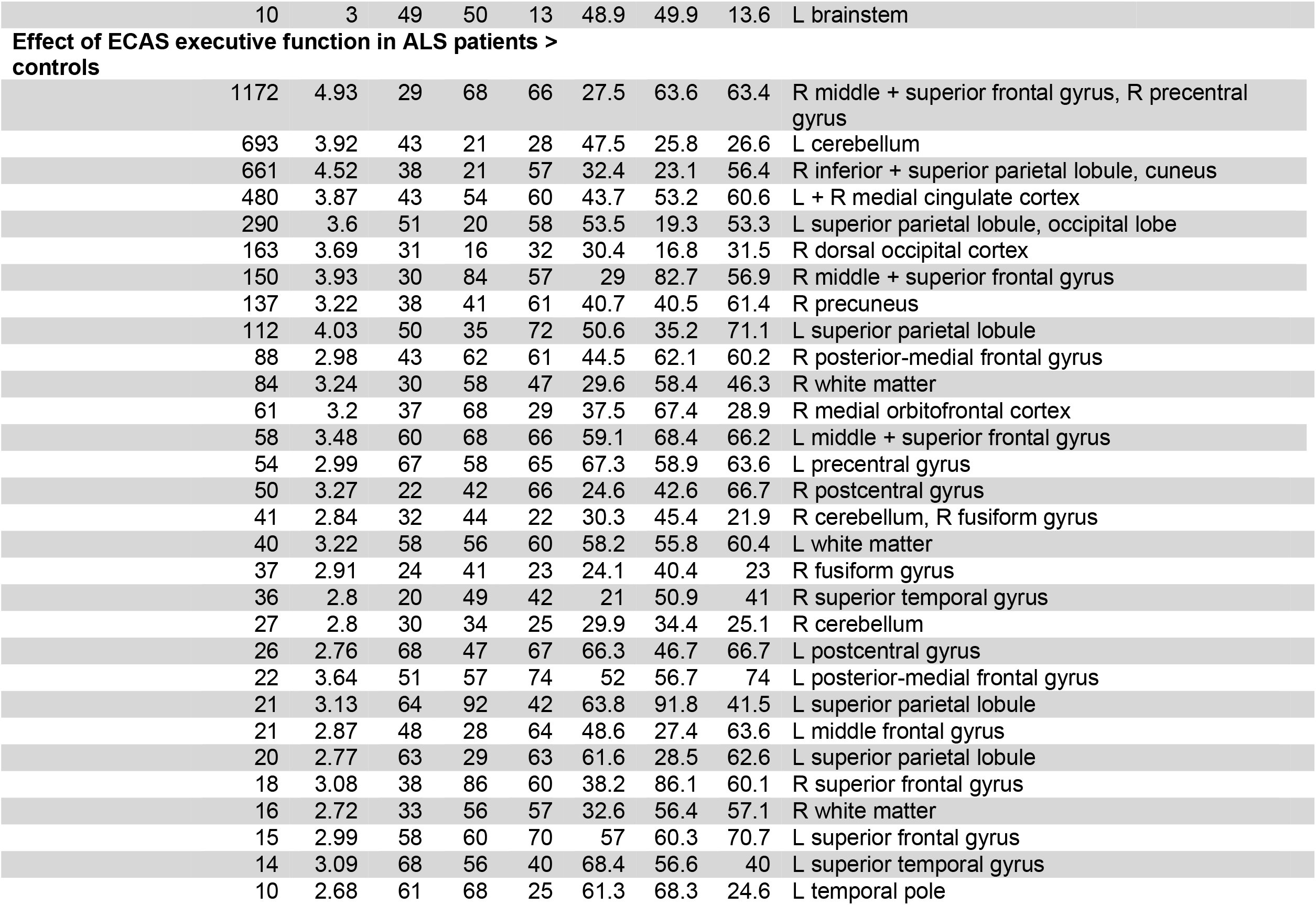
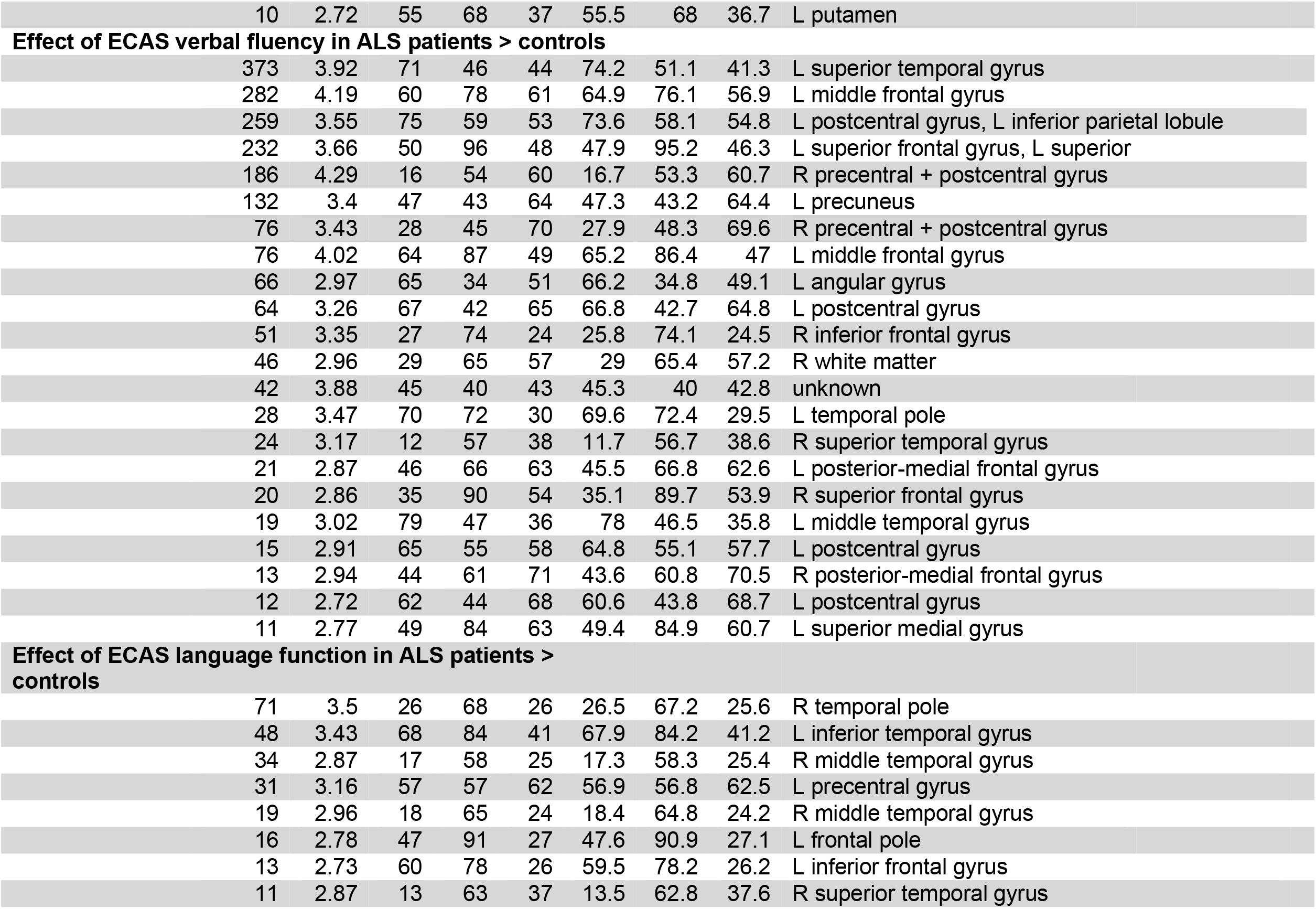
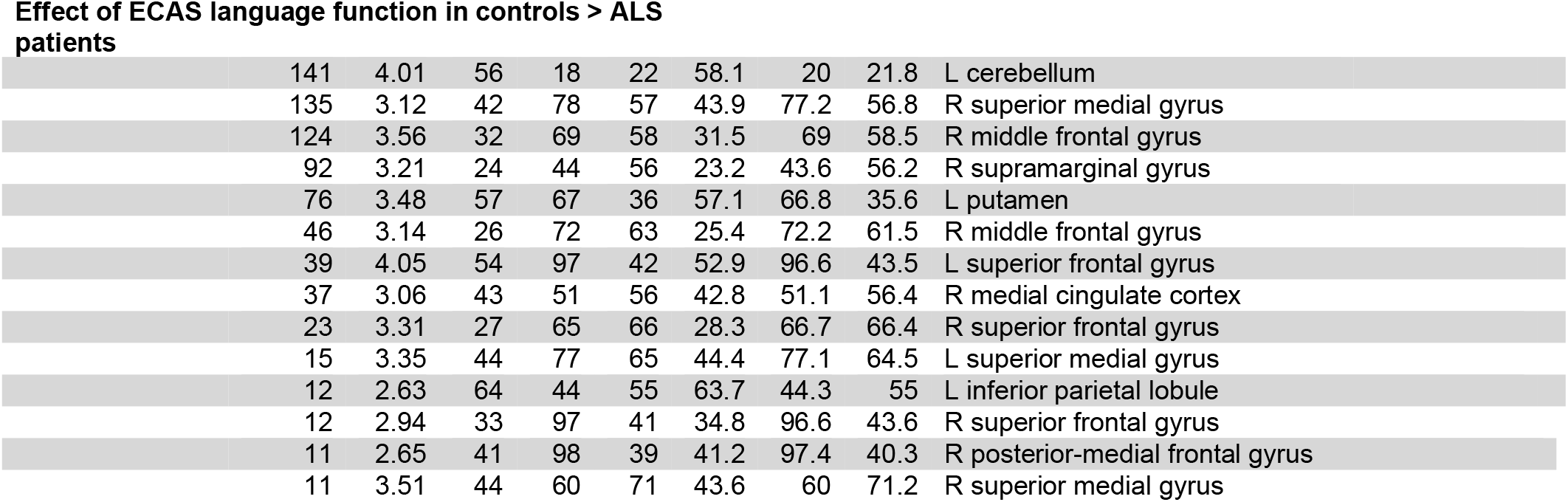
**Regions of significant activity in the 2back > 0-back contrast in controls and ALS patients and group comparison of effects of neuropsychological function on brain activity**.

## References

1. Neumann, M. et al. Ubiquitinated TDP-43 in frontotemporal lobar degeneration and amyotrophic lateral sclerosis. Science 314, 130–133 (2006).

2. DeJesus-Hernandez, M. et al. Expanded GGGGCC hexanucleotide repeat in noncoding region of C9ORF72 causes chromosome 9p-linked FTD and ALS. Neuron 72, 245–256 (2011).

3. Renton, A. E. et al. A hexanucleotide repeat expansion in C9ORF72 is the cause of chromosome 9p21-linked ALS-FTD. Neuron 72, 257–268 (2011).

4. Abrahams, S. et al. Frontotemporal white matter changes in amyotrophic lateral sclerosis. J. Neurol. 252, 321–331 (2005).

5. Mohammadi, B. et al. Amyotrophic lateral sclerosis affects cortical and subcortical activity underlying motor inhibition and action monitoring. Hum. Brain Mapp. 36, 2878–2889 (2015).

6. Stoppel, C. M. et al. Structural and functional hallmarks of amyotrophic lateral sclerosis progression in motor-and memory-related brain regions. NeuroImage Clin. 5, 277–290 (2014).

7. Brettschneider, J. et al. Stages of pTDP-43 pathology in amyotrophic lateral sclerosis. Ann. Neurol. 74, 20–38 (2013).

8. Ringholz, G. M. et al. Prevalence and patterns of cognitive impairment in sporadic ALS. Neurology 65, 586–590 (2005).

9. Montuschi, A. et al. Cognitive correlates in amyotrophic lateral sclerosis: a population-based study in Italy. J. Neurol. Neurosurg. Psychiatry 86, 168–173 (2015).

10. Rippon, G. A. et al. An observational study of cognitive impairment in amyotrophic lateral sclerosis. Arch. Neurol. 63, 345–52 (2006).

11. Phukan, J. et al. The syndrome of cognitive impairment in amyotrophic lateral sclerosis: a population-based study. J. Neurol. Neurosurg. Psychiatry 83, 102–108 (2012).

12. Chiò, A. et al. Cognitive impairment across ALS clinical stages in a population-based cohort. Neurology 93, e984–e994 (2019).

13. Beeldman, E. et al. The cognitive profile of ALS: a systematic review and meta-analysis update. J. Neurol. Neurosurg. Psychiatry 87, 611–9 (2016).

14. Machts, J. et al. Memory deficits in amyotrophic lateral sclerosis are not exclusively caused by executive dysfunction: a comparative neuropsychological study of amnestic mild cognitive impairment. BMC Neurosci. 15, 83 (2014).

15. Girardi, A., Macpherson, S. E. & Abrahams, S. Deficits in emotional and social cognition in amyotrophic lateral sclerosis. Neuropsychology 25, 53–65 (2011).

16. Elamin, M. et al. Executive dysfunction is a negative prognostic indicator in patients with ALS without dementia. Neurology 76, 1263–1269 (2011).

17. Nguyen, C., Caga, J., Mahoney, C. J., Kiernan, M. C. & Huynh, W. Behavioural changes predict poorer survival in amyotrophic lateral sclerosis. Brain Cogn. 150, 105710 (2021).

18. Baddeley, A. D. Working memory. Philos. Trans. R. Soc. B Biol. Sci. 302, 311–324 (1983).

19. Baddeley, A. D. & Hitch, G. Working Memory. Psychol. Learn. Motiv. 8, 47–89 (1974).

20. Baddeley, A. The episodic buffer: a new component of working memory? Trends Cogn. Sci. 4, 417–423 (2000).

21. Repovš, G. & Baddeley, A. The multi-component model of working memory: Explorations in experimental cognitive psychology. Neuroscience 139, 5–21 (2006).

22. Repovš, G. & Baddeley, A. The multi-component model of working memory: Explorations in experimental cognitive psychology. Neuroscience 139, 5–21 (2006).

23. Volpato, C. et al. Working memory in amyotrophic lateral sclerosis: auditory event-related potentials and neuropsychological evidence. J. Clin. Neurophysiol. Off. Publ. Am. Electroencephalogr. Soc. 27, 198–206 (2010).

24. Kasper, E. et al. No change in executive performance in ALS patients: A longitudinal neuropsychological study. Neurodegener. Dis. 16, 184–191 (2016).

25. Hammer, A., Vielhaber, S., Rodriguez-Fornells, A., Mohammadi, B. & Münte, T. F. A neurophysiological analysis of working memory in amyotrophic lateral sclerosis. Brain Res. 1421, 90–99 (2011).

26. Zaehle, T. et al. Working Memory in ALS Patients: Preserved Performance but Marked Changes in Underlying Neuronal Networks. PLoS ONE 8, 1–10 (2013).

27. Vellage, A. K. et al. Working memory network changes in ALS: An fMRI study. Front. Neurosci. 10, 1–10 (2016).

28. Owen, A. M., McMillan, K. M., Laird, A. R. & Bullmore, E. N-back working memory paradigm: A meta-analysis of normative functional neuroimaging studies. Hum. Brain Mapp. 25, 46–59 (2005).

29. Brooks, B. R., Miller, R. G., Swash, M., Munsat, T. L., & World Federation of Neurology Research Group on Motor Neuron Diseases. El Escorial revisited: revised criteria for the diagnosis of amyotrophic lateral sclerosis. Amyotroph. Lateral Scler. Mot. Neuron Disord. Off. Publ. World Fed. Neurol. Res. Group Mot. Neuron Dis. 1, 293–299 (2000).

30. Rascovsky, K. et al. Sensitivity of revised diagnostic criteria for the behavioural variant of frontotemporal dementia. Brain vol. 134 2456–2477 (2011).

31. Cedarbaum, J. M. et al. The ALSFRS-R: A revised ALS functional rating scale that incorporates assessments of respiratory function. J. Neurol. Sci. 169, 13–21 (1999).

32. Abrahams, S., Newton, J., Niven, E., Foley, J. & Bak, T. H. Screening for cognition and behaviour changes in ALS. Amyotroph. Lateral Scler. Front. Degener. 15, 9–14 (2014).

33. Strong, M. J. et al. Amyotrophic lateral sclerosis - frontotemporal spectrum disorder (ALS-FTSD): Revised diagnostic criteria. Amyotroph. Lateral Scler. Front. Degener. 18, 153–174 (2017).

34. Lule, D. et al. The Edinburgh Cognitive and Behavioural Amyotrophic Lateral Sclerosis Screen: a cross-sectional comparison of established screening tools in a German-Swiss population. Amyotroph. Lateral Scler. Front. Degener. 16, 16–23 (2015).

35. Braver, T. S. et al. A Parametric Study of Prefrontal Cortex Involvement in Human Working Memory. NeuroImage 5, 49–62 (1997).

36. The jamovi project. jamovi (Version 1.2) (2020).

37. Jenkinson, M., Bannister, P., Brady, M. & Smith, S. Improved optimization for the robust and accurate linear registration and motion correction of brain images. NeuroImage 17, 825–841 (2002).

38. Smith, S. M. Fast robust automated brain extraction. Hum. Brain Mapp. 17, 143–155 (2002).

39. Beckmann, C. F. & Smith, S. M. Probabilistic independent component analysis for functional magnetic resonance imaging. IEEE Trans. Med. Imaging 23, 137–152 (2004).

40. Abraham, A. et al. Machine learning for neuroimaging with scikit-learn. Front. Neuroinformatics 8, (2014).

41. Rottschy, C. et al. Modelling neural correlates of working memory: A coordinate-based meta-analysis. Neuroimage 60, 830–846 (2012).

42. Emch, M., von Bastian, C. C. & Koch, K. Neural Correlates of Verbal Working Memory: An fMRI Meta-Analysis. Front. Hum. Neurosci. 13, (2019).

43. Charroud, C. et al. Working memory activation of neural networks in the elderly as a function of information processing phase and task complexity. Neurobiol. Learn. Mem. 125, 211–223 (2015).

44. Demeter, E., Hernandez-Garcia, L., Sarter, M. & Lustig, C. Challenges to attention: A continuous arterial spin labeling (ASL) study of the effects of distraction on sustained attention. NeuroImage 54, 1518–1529 (2011).

45. Engstrom, M., Landtblom, A.-M. & Karlsson, T. Brain and effort: brain activation and effort-related working memory in healthy participants and patients with working memory deficits. Front. Hum. Neurosci. 7, (2013).

46. Jiménez-Balado, J. & Eich, T. S. GABAergic dysfunction, neural network hyperactivity and memory impairments in human aging and Alzheimer’s disease. Semin. Cell Dev. Biol. (2021) doi:10.1016/j.semcdb.2021.01.005.

47. Attwell, D. & Laughlin, S. B. An energy budget for signaling in the grey matter of the brain. J. Cereb. Blood Flow Metab. Off. J. Int. Soc. Cereb. Blood Flow Metab. 21, 1133–1145 (2001).

48. Vandoorne, T., De Bock, K. & Van Den Bosch, L. Energy metabolism in ALS: an underappreciated opportunity? Acta Neuropathol. (Berl.) 135, 489–509 (2018).

49. Keller, J. et al. Functional reorganization during cognitive function tasks in patients with amyotrophic lateral sclerosis. Brain Imaging Behav. (2017) doi:10.1007/s11682-017-9738-3.

50. Schoenfeld, M. A. et al. Functional motor compensation in amyotrophic lateral sclerosis. J. Neurol. 252, 944–952 (2005).

51. Witiuk, K. et al. Cognitive Deterioration and Functional Compensation in ALS Measured with fMRI Using an Inhibitory Task. J. Neurosci. 34, 14260–14271 (2014).

52. Scheller, E., Minkova, L., Leitner, M. & Klöppel, S. Attempted and Successful Compensation in Preclinical and Early Manifest Neurodegeneration –A Review of Task fMRI Studies. Front. Psychiatry 5, (2014).

53. Cabeza, R., Dennis, N., Stuss, D. T. & Knight, R. T. Principles of frontal lobe function. Ed 2, 628–652 (2012).

54. Abrahams, S. et al. Verbal fluency and executive dysfunction in amyotrophic lateral sclerosis (ALS). Neuropsychologia 38, 734–747 (2000).

55. Evdokimidis, I. et al. Frontal lobe dysfunction in amyotrophic lateral sclerosis. J. Neurol. Sci. 195, 25–33 (2002).

56. Meier, S. L., Charleston, A. J. & Tippett, L. J. Cognitive and behavioural deficits associated with the orbitomedial prefrontal cortex in amyotrophic lateral sclerosis. Brain J. Neurol. 133, 3444–3457 (2010).

57. Gregory, J. M. et al. Executive, language and fluency dysfunction are markers of localised TDP-43 cerebral pathology in non-demented ALS. J. Neurol. Neurosurg. Psychiatry 91, 149–157 (2020).

58. Trojsi, F. et al. Microstructural Changes across Different Clinical Milestones of Disease in Amyotrophic Lateral Sclerosis. PloS One 10, e0119045 (2015).

59. Sorrentino, P. et al. Flexible brain dynamics underpins complex behaviours as observed in Parkinson’s disease. Sci. Rep. 11, 4051 (2021).

60. Wager, T. D., Lindquist, M. & Kaplan, L. Meta-analysis of functional neuroimaging data: current and future directions. Soc. Cogn. Affect. Neurosci. 2, 150–158 (2007).

61. Nyberg, L. et al. Longitudinal evidence for diminished frontal cortex function in aging. Proc. Natl. Acad. Sci. U. S. A. 107, 22682–22686 (2010).

62. Poletti, B. et al. The Arrows and Colors Cognitive Test (ACCT): A new verbal-motor free cognitive measure for executive functions in ALS. PloS One 13, e0200953 (2018).

63. Keller, J. et al. A first approach to a neuropsychological screening tool using eye-tracking for bedside cognitive testing based on the Edinburgh Cognitive and Behavioural ALS Screen. Amyotroph. Lateral Scler. Front. Degener. 18, 443–450 (2017).

64. Steinbach, R., Gaur, N., Stubendorff, B., Witte, O. W. & Grosskreutz, J. Developing a Neuroimaging Biomarker for Amyotrophic Lateral Sclerosis: Multi-Center Data Sharing and the Road to a “Global Cohort”. Front. Neurol. 9, (2018).

65. Kasper, E. et al. No change in executive performance in ALS patients: A longitudinal neuropsychological study. Neurodegener. Dis. 16, 184–191 (2016).

